# Effectiveness of bimodal neuromodulation for tinnitus treatment in a real-world clinical setting in United States: A retrospective chart review

**DOI:** 10.1101/2024.08.22.24312175

**Authors:** Emily E. McMahan, Hubert H. Lim

## Abstract

Bimodal neuromodulation combining sound therapy with electrical tongue stimulation using the Lenire device is emerging as an effective treatment for tinnitus. A single-arm retrospective chart review analyzes real-world outcomes for 220 tinnitus patients from the Alaska Hearing and Tinnitus Center for the recently FDA-approved Lenire treatment for the first time in a United States clinic. The primary endpoint examines the responder rate and mean change in Tinnitus Handicap Inventory (THI) after approximately 12 weeks of treatment in eligible patients with moderate or worse tinnitus. A responder represents a THI improvement of greater than seven points (i.e., minimal clinically important difference, MCID). Of 212 patients with available data, there was a high responder rate of 91.5% (95% CI: 86.9%, 94.5%) with a mean improvement of 27.8 ± 1.3 (SEM) points, and no device-related serious adverse events. Furthermore, a THI MCID of seven points represents a consistent criterion for clinical benefit based on real-world evidence.

## Introduction

Tinnitus, the phantom auditory experience without an external source affects 10-15% of the global population^1–6^. For some, tinnitus is a minor annoyance while for approximately 6-11% of the tinnitus population, the experience is bothersome^3^. Unfortunately, there are limited treatment options for bothersome tinnitus^7–9^. When left unmanaged, bothersome tinnitus can be debilitating with a significant negative impact on the patient’s quality of life^7–9^. In the United States, tinnitus was the most common service-connected disability claimed by 2.9 million veterans in 2023^10^. Overall, tinnitus remains a major health issue in our society.

One promising noninvasive and accessible treatment approach supported by animal studies^11^ and several large-scale clinical trials is bimodal neuromodulation^12–14^, which combines sound therapy with electrical tongue stimulation using the Lenire device (Neuromod Devices, Ireland; Fig. 1a). More recently in March 2023, Lenire was granted De Novo approval for the treatment of tinnitus by the Food and Drug Administration (FDA; DEN210033)^12^. Results from the controlled pivotal clinical trial that was designed with guidance from the FDA confirmed that for those with moderate or more severe tinnitus symptoms (Tinnitus Handicap Inventory, THI, greater than or equal to 38) when starting bimodal treatment, a clinically significant superior performance of bimodal neuromodulation (i.e., improvement greater than seven points on THI) was achieved with just six weeks of treatment compared to sound-only stimulation^12^. The 7-points improvement in THI is a criterion for the minimal clinically important difference (MCID) that has been estimated with a combination of an anchor-based method (i.e., CGI-I) and distribution-based method (i.e., effect size of 0.5 of SD)^15^, consistent with FDA guidelines for determining a threshold for significant change^16^.

**Fig. 1.**
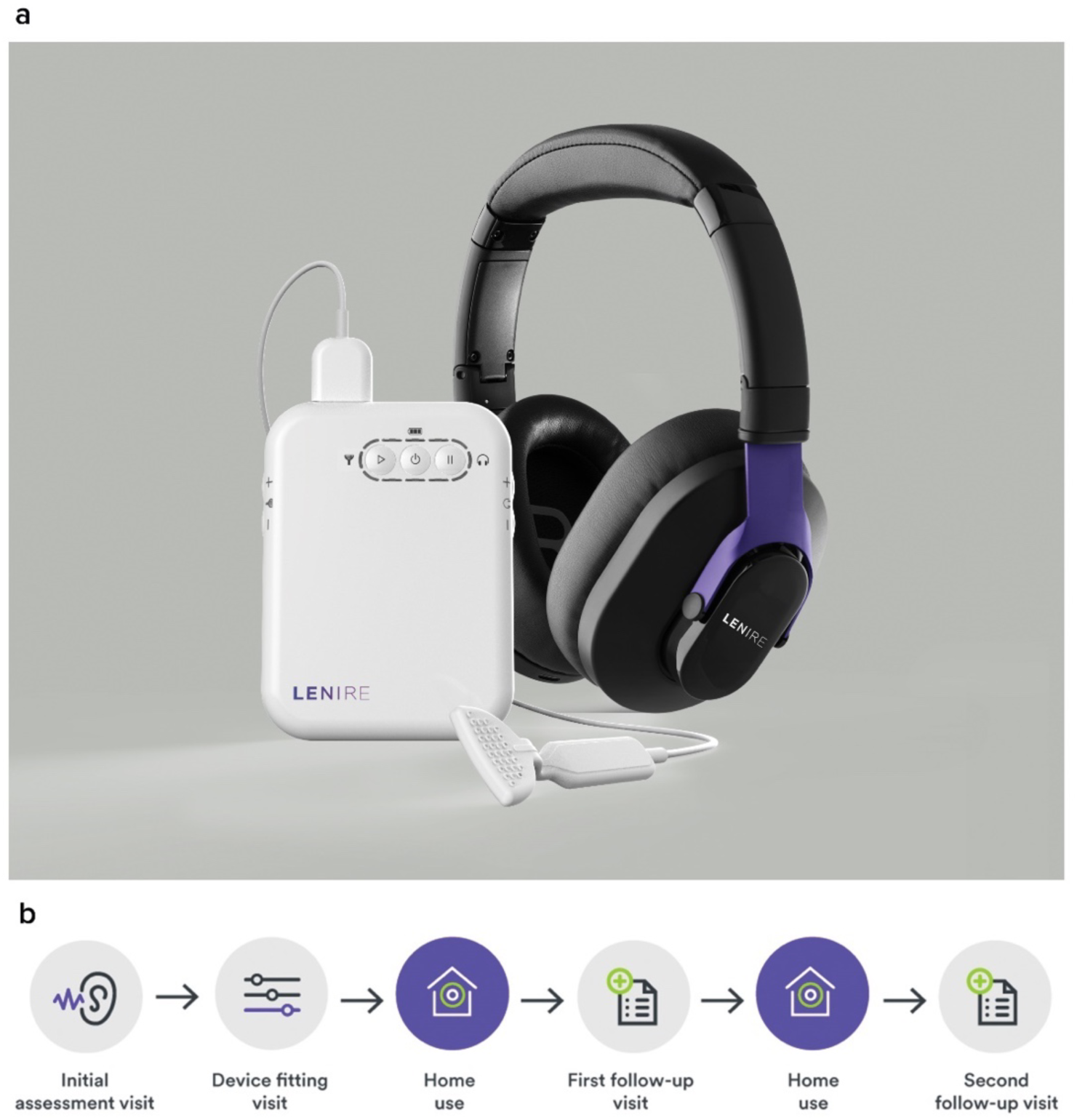
**(a)** Lenire bimodal neuromodulation device by Neuromod Devices (Dublin, Ireland) intended to reduce the symptoms of tinnitus in patients with moderate or worse tinnitus (i.e., FDA-approved for those with a Tinnitus Handicap Inventory (THI) score greater than or equal to 38). The device consists of a Tonguetip®, an intraoral device designed to sit comfortably in the mouth and deliver gentle electrical stimulation on the tongue’s surface; Bluetooth headphones that play personalized sounds to the ears; and a handheld controller for patients to adjust the duration and intensity of the treatment. The patient has a limited range of control of the sound volume and tongue stimulation with the handheld controller for ease of comfort and to maintain stimulation sensations at a noticeable but near threshold levels. **(b)** Lenire standard of care procedure at Alaska Hearing and Tinnitus Center.

Although the controlled pivotal trial led to positive results for tinnitus treatment, there remains the critical question of how Lenire will perform in a real-world clinical setting that is less structured with greater heterogeneity of patients than in a clinical study. After FDA approval, there were 14 clinics in the United States who were the first group of providers to treat tinnitus patients with the Lenire device. The largest number of patients across those clinics have been treated at the Alaska Hearing and Tinnitus Center with already 220 patients fitted with the Lenire device that is the first to be available for publication as a single site, single arm retrospective chart review. At most of these clinical providers, tinnitus management conventionally involves in-person care; however, since the COVID-19 pandemic, telehealth has become instrumental for routine follow-ups and consultations^17^. Thus, at the Alaska Hearing and Tinnitus Center, tinnitus patients were treated with a hybrid model where Lenire device fitting occurred in-person with follow-ups performed virtually for most patients.

After initial consultation (in-person or via telehealth), if a patient was prescribed Lenire, an in-person device fitting was completed. During in-patient fitting, electrical tongue stimulus intensity was calibrated to a comfortable sensation level for the patient and the sound stimulus was adjusted to a comfortable loudness based on the audiogram for each patient. All patients utilized the same stimulation setting, which includes pure tones presented to the ears that are synchronized with electrical pulses presented to the top surface of the tongue (further details provided in previous publications)^12,13,18^. Patients were provided with the User Manual and comprehensive training with the device, including what to expect from the treatment, potential side effects, and how to use the device. Patients were instructed to use the device for up to 60 minutes per day for at least 12 weeks (Fig. 1b). Follow-on care and assessments were carried out approximately halfway through their treatment (FU1) and approximately 12 weeks (FU2) after initial assessment; these were completed via telehealth with the option for in-person care for those who were located closer to a clinic. The online services facilitate continued care by providing the possibility for remote tinnitus counselling, education, and additional consultations. This observational study was reviewed before initiation by a registered IRB (IRB number 00000971; Columbia, Maryland, USA; study number Pro00077817). The IRB determined the research project was exempt from IRB oversight under 45 CFR 46.104(d) (4).

## Results

### Characteristics of patients

There were 220 patients who satisfied the FDA labelling criterion of a THI score ≥ 38 and were fitted with the Lenire bimodal neuromodulation device at the Alaska Hearing and Tinnitus Center clinics from May 4, 2023 to March 28 2024 (Fig. 2). The demographic characteristics of these patients are listed in Table 1. The mean age of patients was 60.3 ± 12.6 years with a mean tinnitus duration of 8.5 ± 10.0 years. Of the 220 patients, 73.2% were males and 26.8% were females, which is consistent with the prevalence of tinnitus in the literature being higher in males than females across demographic groups^5,19,20^.

**Fig. 2.**
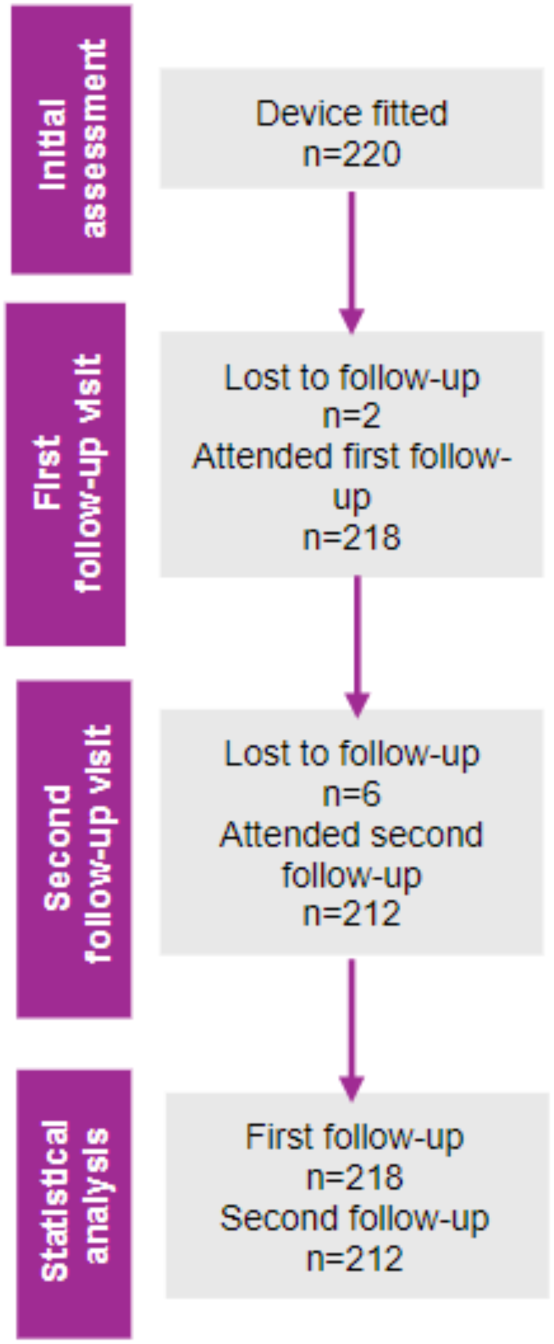
Patient flow diagram. There were 220 patients with moderate or worse tinnitus severity (THI ≥ 38) who were fitted with the Lenire device at initial assessment in accordance with FDA device labeling.

**Table 1.** Demographics and tinnitus characteristics of all available patient data (n=220) at initial assessment.

| **All available patients (n=220)** |  |
| --- | --- |
| **Age (years)**<br>Mean ± SD (n) | 60.3 ± 12.6 (220) |
| **Sex [% (n/N)]**<br>Male<br>Female | 73.2% (161/220)<br>26.8% (59/220) |
| **Tinnitus duration at initial assessment (years)**<br>Mean ± SD (n) | 8.5 ± 10.0 (220) |
| **THI at initial assessment (points)**<br>Mean ± SD (n) | 60.0 ± 17.4 (220) |
| **Hearing loss at initial assessment (dB HL)**<br>Mean ± SD (n)<br>Right ear<br>Left ear | 19.6 ± 12.4 (215)<br>20.4 ± 13.5 (214) |

Mean hearing loss at initial assessment was calculated using the average of 500 Hz, 1000 Hz and 2000 Hz for each ear. THI: Tinnitus Handicap Inventory; dB HL: decibel hearing level.

At FU1, there were only two patients who were lost to follow-up, with six patients who were lost to follow-up at FU2 (Fig. 2), resulting in a high retention rate of 96.4%. There were consistent characteristics of patients who were lost to follow up relative to the full cohort (see Supplementary Table 1). For analysis, there were data for 218 patients at FU1 and 212 patients at FU2.

### Clinical efficacy and safety of bimodal treatment are replicable in the real-world

Confirming the benefit of the Lenire treatment for tinnitus observed in previous large-scale clinical studies^12–14^, our primary endpoint analysis demonstrated that 91.5% (95% CI: 86.9%, 94.5 %) of patients with moderate or worse tinnitus severity achieved clinically significant benefit exceeding MCID after approximately 12 weeks of treatment (Fig. 3a), corresponding to 27.8 ± 1.3 (SEM) points reduction in tinnitus severity (Fig. 3b). Encouragingly, even after approximately six weeks of treatment by FU1 (i.e. only halfway through the recommended treatment plan), 78.0% (95% CI: 72.0%, 83.0%) of patients already achieved clinically significant benefit (Fig. 3a), corresponding to 18.5 ± 1.1 (SEM) points reduction in tinnitus severity (Fig. 3b). For completeness, the mean changes in THI from initial assessment to FU1 and FU2 for patients who returned at both follow-up visits are shown in Fig. 3c, depicting additional improvement in tinnitus symptoms over time with continued treatment. As shown across different types of analyses in Fig. 3, there is a significant improvement in tinnitus symptoms over time achieved with continued use of the Lenire treatment. Clinical efficacy results in terms of responder rate and mean changes in THI are consistently observed for male and female participants (Supplementary Fig. 1), with responses for individual patients shown as scatter plots in Supplementary Fig. 2.

**Fig. 3.**
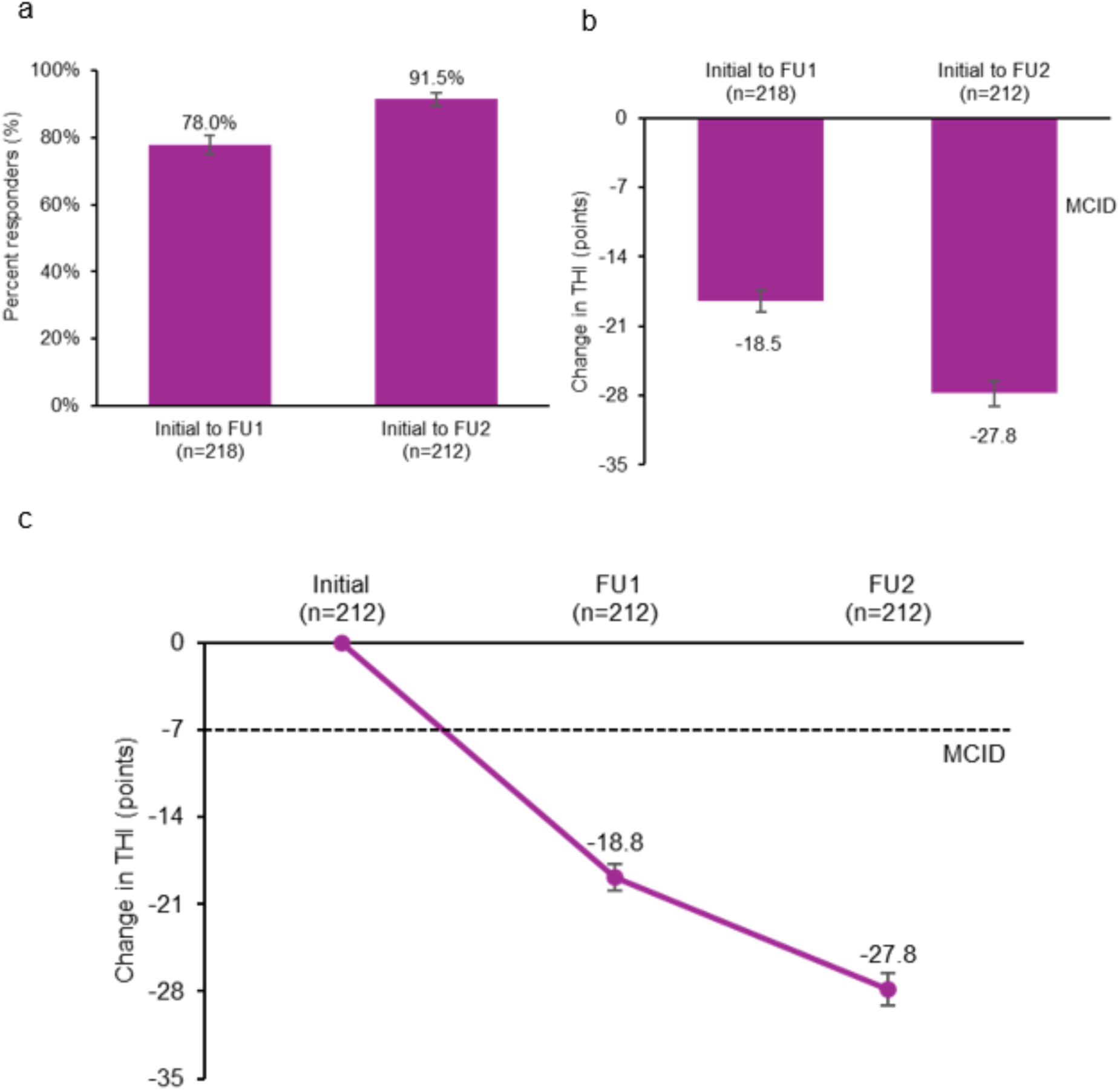
**(a)** Percent responders (MCID improvement in THI > 7 points) from initial assessment to first follow-up (FU1) and second follow-up (FU2); SEM bars are shown. Two-sided Z-test of proportions for comparison between groups; p<0.001. **(b)** Mean improvement in THI score from initial assessment to FU1 and FU2; SEM bars are shown. Independent t-test for comparison between groups; p<0.001. **(c)** Mean change in THI from initial assessment to FU1 and FU2 for patients who returned at both follow-up visits; SEM bars are shown. Paired t-test for comparison between groups from initial assessment to FU1 versus to FU2; p<0.001. MCID; minimal clinically important difference. THI; Tinnitus Handicap Inventory.

Across 220 patients, there were only eight patients that were lost to follow up by FU2. Since the patients paid out of pocket ($4500) for the Lenire treatment, it is expected that if the patients were not sufficiently improving in their symptoms, they would return back to the clinic for further clinical support. Thus, it is assumed that a majority or all of those eight patients were responders. This scenario appears to be the case, by analyzing the change in THI score for at least six of the eight patients who still came to FU1 where five of the six already had improvements in THI scores after 6 weeks of treatment (see Supplementary Table 2). By FU2, these five patients and possibly the sixth patient are assumed to have achieved enough improvements to opt out of returning for FU2. The other two patients who did not show up to FU1 may also have improved sufficiently with 6 weeks of treatment to opt not to return to any follow-up visits. Even assuming that all eight of those lost to follow-up patients are non-responders, the responder rate would still be a high rate of 88.2% (194 out of 220).

In terms of safety, there have been no device related serious adverse events (SAEs) or medical field experience events that warranted reporting to the manufacturer or FDA that were outside the normal issues experienced at the Alaska Hearing and Tinnitus Center during the standard care of tinnitus patients; thus, the Lenire treatment has continued to exhibit a high benefit to safety profile as observed in the FDA pivotal trial^12^.

### High self-reported benefit rate is consistent with 7-points MCID for THI

At FU2, we directly asked patients if they found Lenire to be beneficial for their tinnitus journey, in which a high percent of 89.2% indicated yes (Fig. 4a). It is noteworthy that the patients who reported that they benefitted from Lenire were aligned well with those who improved by at least the MCID cut-off of 7 points in their THI score (Fig. 4a), consistent with previous research that defined the MCID for THI^15^. At a more conservative cut-off of 9 points (i.e., THI ≥ 10), there would be a more equivalent number of patients who benefitted from treatment that would be at or below that cut-off (seven missed responders) versus those who did not benefit who would be at or above that cut-off (six false responders). This conservative MCID of 9 points still leads to a high responder rate of 88.7% (95% CI; 83.7%, 92.3 %) by FU2 (Fig. 4b). Therefore, a MCID of seven points is a clinically consistent criterion for representing a minimal meaningful benefit in tinnitus symptoms, with an upper conservative criterion of 9 points, and is based on real-world evidence with a large patient cohort treated with the Lenire treatment.

**Fig. 4.**
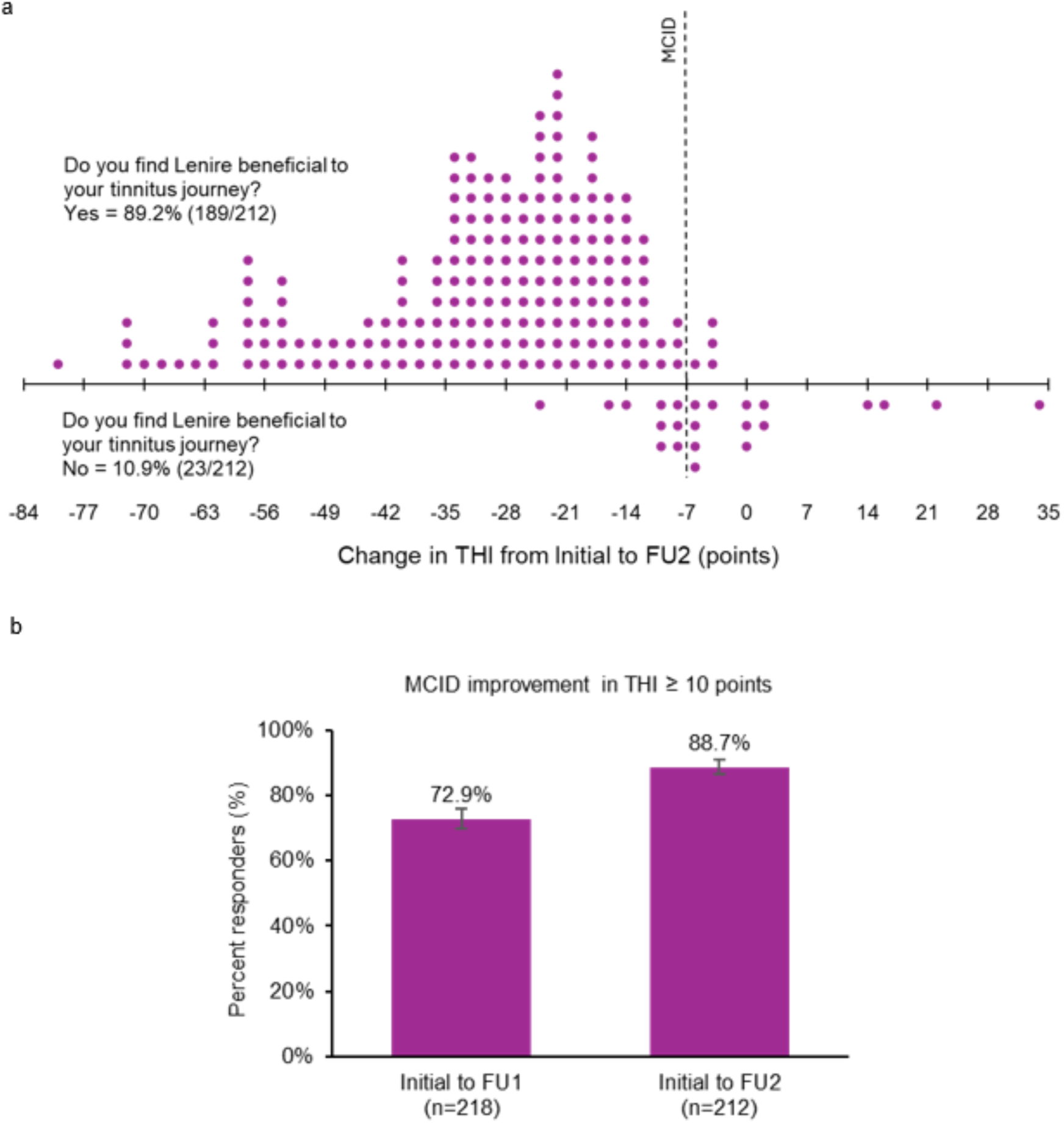
**(a)** Patient reported benefit in using the Lenire device for treating tinnitus in relation to their improvement in THI score, demonstrating that benefit is well aligned with a MCID cut-off of seven points. **(b)** Percent responders from initial assessment to FU1 and FU2 using a more conservative MCID of 9 points (i.e., greater than or equal to 10 points) where there is a more equivalent number of misses versus false hits (i.e., seven patients in the upper-right quadrant versus six patients in the lower-left quadrant, respectively). MCID: minimal clinically important difference; FU1: first follow-up visit; FU2: second follow-up visit.

## Discussion

The main objective of this retrospective chart review was to confirm the clinical efficacy and safety of the Lenire bimodal treatment in a real-world clinical setting in the United States, since obtaining FDA approval for the treatment in March of 2023. The indications for use for Lenire are to alleviate symptoms of tinnitus in patients 18 years of age and older suffering from at least moderate severity tinnitus (THI ≥ 38 at initial assessment). In this patient population, the results were positive and consistent with previous large scale clinical trials and initial real-world evidence published from Europe ^12–14,21^, demonstrating that 91.5% of patients exceeded the MCID after approximately 12 weeks of treatment. In a worst-case scenario using a more conservative MCID of 9 points, there was still a high responder rate of 88.7%; or even assuming the eight lost to follow-up patients were all non-responders (out of 220 patients), then the responder rate would still be at 88.2%. Directly asking patients if they benefitted from the Lenire treatment also led to a high success rate of 89.2%. In terms of safety, there were no serious or major device related adverse events reportable to the manufacturer or FDA from the Alaska Hearing and Tinnitus Center. As shown in Fig. 4a, there were four patients who exhibited noticeable increases in their THI scores who indicated that they did not benefit from the treatment by the 12-week assessment. These patients were further supported with CBT, counseling, breathing exercises, and other habituation methods to help with their tinnitus. All but one patient has returned to their baseline situation prior to starting Lenire treatment. The one patient who has not returned to their baseline has been experiencing a tragic personal and health event not related to Lenire treatment that is likely contributing to their tinnitus status. Overall, as observed in the FDA pivotal trial and previous large-scale clinical studies^12–14^, the real-world clinical data supports a high benefit to risk profile for the Lenire treatment.

Encouragingly, the proportion of patients who benefitted from treatment was well aligned with those who received a clinically meaningful improvement from the Lenire intervention as assessed with the validated THI^15^. A MCID of seven points for THI has been demonstrated to be a valid clinically relevant criterion for assessing meaningful benefit from an intervention, based on our real-world clinical data with the Lenire treatment in a large patient cohort.

Another objective of this study was to assess the feasibility of a hybrid tinnitus management approach for the Lenire treatment. Telehealth encompasses different modalities and is used for diverse health conditions and patient populations^22^. In tinnitus, to decrease the burden of repeated in-person care, treatment delivered via telehealth are on the rise^17^. Since March 2020 associated with the COVID pandemic, both hybrid and fully remote telehealth services for tinnitus patients have become more mainstream with benefits shown throughout the therapeutic process^17^. However, it has been highlighted in a systematic review of telehealth interventions for tinnitus (iCBT, internet-based interventions, self-help devices, and smartphone apps) that the main barriers to the success are due to a high dropout rate and lack of adherence to treatment^17^. Encouragingly, our high responder rates along with the high satisfaction and retention rates indicate that patients had a positive and effective experience with our hybrid delivery model of tinnitus care for bimodal treatment that incorporates telehealth and in-person services. Such a hybrid model greatly opens up the opportunity to treat a much larger patient population suffering from tinnitus in an accessible and scalable way.

Tinnitus is a well-known heterogenous disorder that requires experienced clinicians with a deep knowledge of methods and technologies available to help the patient manage their disturbing condition. These results are the first real-world evidence for Lenire tinnitus treatment published from a United States cohort and provides confirmation that the treatment efficacy of the Lenire device can be successfully translated and replicated into a clinical practice setting. When new treatment devices enter clinics and patient care settings, the benefits can be lower than observed in structured clinical studies due to a greater diversity of patients and variability in clinical processes in the real-world environment. Therefore, it is important to continue tracking the effectiveness of novel treatments in the real-world setting to build evidence-based care pathways for tinnitus patients. Our patient results combined with previous clinical studies^12–14^ further support the Lenire device as an effective and safe treatment for tinnitus.

## Methods

### Study design

This study is a single site, single arm retrospective analysis of 220 patients who were fitted with the Lenire device from May 4, 2023 to March 28, 2024 at the Alaska Hearing and Tinnitus Center clinics. The patients were fitted based on the indications for use for Lenire in patients 18 years of age and older suffering from at least moderate tinnitus severity (THI ≥ 38 at initial assessment). This observational study was reviewed by a registered IRB (IRB number: 00000971; Study Protocol number: Pro00077817) and was determined to be exempt from IRB oversight based on the Department of Health and Human Services regulations found at 45 CFR 46.104(d)(4). All methods were carried out in accordance with relevant guidelines and regulations, in which the study protocol is available in the Supplementary Information.

Medical records stored in CounselEAR Office Management Solutions (OMS) were accessed by the clinician who had access to the records as part of her standard duties at the clinic. Upon exporting the data, no identifiable information was retained in the final study database, nor will there be a need to re-identify the data. Patients in this study will not be contacted and have given written consent to a HIPAA Waiver Notice (‘Notice of Privacy Practices’) prior to initial treatment at the clinic, which allows for data to be used for research purposes. It was verbally explained during the initial assessment how the data would be used to further research on the Lenire device. A template of the clinic’s Privacy Policy and Notice of Privacy Practices was attached to the IRB submission as supporting information.

All patients in this study attended an initial assessment via CounselEAR’s telehealth portal or in clinic, where health evaluations and several tinnitus assessments were performed. All patients received tinnitus counseling and education prior to selecting a treatment plan that best supports their hearing and tinnitus needs. The audiologist reviewed all appropriate expectations on outcomes and commitments with each patient in terms of treatment compliancy. Patients were fitted with the Lenire device and were provided with a device training session and the Lenire User Manual. During fitting, electrical tongue stimulus intensity was calibrated to a comfortable sensation level and sound stimulus was adjusted to a comfortable loudness based on the audiogram for each patient. They were instructed to use the device for up to 60 minutes per day for at least 12 weeks. Severity of tinnitus was assessed using the THI at FU1 approximately halfway through their treatment, and at a second follow-up approximately 12 weeks from initial fitting (FU2). Patients could contact the clinic at any time between appointments if there were any concerns. Virtual video calls were primarily used for follow-up assessments, additional consultations, counselling, and education.

The Lenire neuromodulation system is an FDA approved take-home device. Treatment sessions were self-administered by patients at home. A detailed description of the device has been published in previous Lenire clinical trials^12–14^. All patients in this study utilized the same stimulation setting (PS1), which includes pure tones presented to the ears that are synchronized with electrical pulses presented to the top surface of the tongue. There are currently two stimulation settings, PS1 and PS6, which are approved by FDA for tinnitus treatment in the United States based on the results from the TENT-A3 pivotal trial and supporting clinical data included in the FDA de novo submission. A full description of the PS1 and PS6 settings can be found in previous clinical trial publications^13,14^.

### Participants

Lenire is prescribed to patients 18 years and older with subjective tinnitus that is moderate or worse in terms of tinnitus severity (THI ≥ 38). The device is contraindicated for those who have an active implantable device; are pregnant; have epilepsy or other conditions which may cause loss of consciousness; have conditions that cause impaired sensitivity in the tongue; have lesions, sores, or inflammation of the oral cavity; have any intermittent or chronic neuralgia in the head and neck area; have Meniere’s disease; have objective tinnitus; and cannot remove oral piercings during device use.

### Clinical endpoints

One of the main objectives of this retrospective chart review is to assess real-world data to determine the replicability of Lenire treatment outcomes observed in previous clinical trials for the first time in a clinical setting within the United States. Therefore, in line with previous clinical trials, THI was used at all stages to assess tinnitus symptom severity for comparability to existing published results^12–14^.

The THI is a validated 25-item questionnaire measuring perceived tinnitus handicap severity anchored with “No” (0 points), “Sometimes” (2 points) or “Yes” (4 points) responses^23^. These scores are added up to a total value ranging from 0 to 100, where a higher score indicates a higher level of tinnitus severity. The THI can also be divided into five severity categories: slight (0-16 points), mild (18-36 points), moderate (38-56 points), severe (58-76 points), and catastrophic (78-100 points). The MCID reported for THI is seven points and represents a clinically meaningful change in tinnitus symptoms^15^.

In addition to the THI, patients were asked at FU2 to respond ‘Yes’ or ‘No’ to the question: “Do you find Lenire beneficial to your tinnitus journey?”

### Field safety reporting

The Lenire device was approved for distribution in the United States in March of 2023 by the FDA. All healthcare professionals providing the Lenire device may submit information in relation to adverse events (AEs), technical issues or general feedback directly to the manufacturer (Neuromod Devices) through the Zoho ticketing system. Once a ticket is received, it is assessed for alleged device malfunction, device fitting problems, undesired product performance or other adverse field experiences (e.g., undesirable medical symptoms), after which it is determined as to the need for a Field Product Experience Report (FPER) to be opened to further document and investigate the report. Once an FPER is opened, an initial determination of the need to submit a report to FDA (in United States) or Competent Authorities (outside the United States) is conducted. Reports are assessed by the clinical or technical teams at the manufacturing company, as appropriate, for the need for further information or follow-up. Where a FPER is determined to warrant reporting to the FDA or a Competent Authority, this is completed using the process and timeline for the applicable jurisdiction. There were no medical FPERs submitted to the manufacturer, and all technical queries were resolvable with no reporting actions required.

### Statistical analyses

The primary endpoint consisted of a responder rate analysis where the responder rate was calculated as the percentage of participants achieving more than seven points reduction in THI from initial assessment to FU2. Responder rate is reported with corresponding 95% CIs. The primary endpoint also included the mean change in symptoms of tinnitus based on THI from initial assessment to FU2 that is reported with corresponding SEM. Additional analyses are reported for responder rate and mean change from initial assessment to FU1, as well as using a more conservative MCID criterion of 9 points based on new findings observed in this study.

To assess differences in performance between follow-up visits, a two-sided Z-test of proportions on responder rates and an independent t-test on mean changes were performed for the THI values. In addition, a paired t-test was conducted to assess the significance between follow-up visits in mean changes in THI for patients who returned for both follow-up visits. At the initial visit, participants self-reported if they were male or female. Analyses were further carried out according to participants’ self-reported sex. Of the 220 participants, 161 were male and 59 were female. All statistical analyses were conducted with Stata 15. Bonferroni correction for multiple comparisons still led to statistically significant results since all tests showed highly statistically significant p-values less than 0.001.

## Data Availability

All relevant data associated with the published study are present in the paper or the Supplementary Information. Access to the raw individual level data may be obtained, contingent on appropriate ethics approval and data sharing agreements, by contacting EEM for the purposes of confirming the analysis in the paper. Responses to valid requests will be reasonably attempted and initiated within 10 working days of receipt beginning 3 months and ending 5 years after this article publication.

## Acknowledgements

The authors would like to thank the team at the Alaska Hearing and Tinnitus Center for their assistance on project and clinic administration, as well as data collection, validation and analyses. The authors also appreciate the technical assistance and administrative support provided by S.L. Leong and E. Meade at Neuromod Devices for this study. Finally, the authors thank the IRB for their review and determination that the study meets the requirements of 45 CFR 46.104(d)(4). All methods were carried out in accordance with relevant guidelines and regulations, in which the study protocol is available in the Supplementary Information.

## Author contributions statement

Conceptualization and methodology: EEM and HHL

Investigation: EEM

Project administration: EEM

Data validation and analyses: EEM and HHL

Writing and editing: EEM and HHL

## Supplementary Information

**Supplementary Figure 1.**
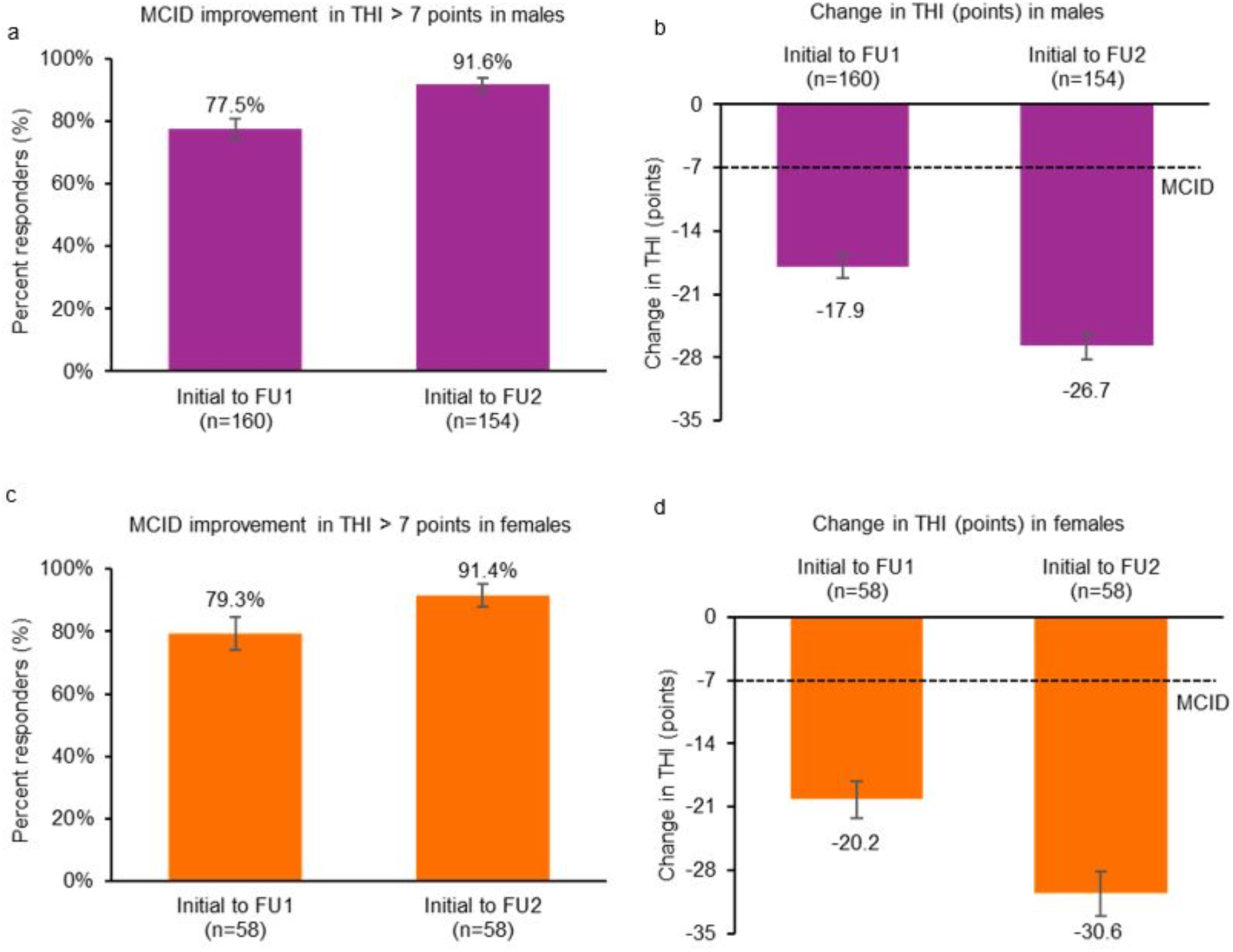
**(a)** In males: percent responders (MCID improvement in THI > 7 points) from initial assessment to first follow-up (FU1) and second follow-up (FU2); SEM bars are shown. Two-sided Z-test of proportions for comparison between groups; p<0.001**. (b)** In males: mean improvement in THI score from initial assessment to FU1 and FU2; SEM bars are shown. Independent t-test for comparison between groups; p<0.001. **(c)** In females: percent responders (MCID improvement in THI > 7 points) from initial assessment to FU1 and FU2; SEM bars are shown. One sample test of proportions for comparison between groups; p=0.001. **(d)** In females: mean improvement in THI score from initial assessment to FU1 and FU2; SEM bars are shown. Paired t-test for comparison between groups; p<0.001. All data supports a significant improvement in tinnitus symptoms over time with continued use of the Lenire treatment that is consistently observed across males and females.

**Supplementary Figure 2.**
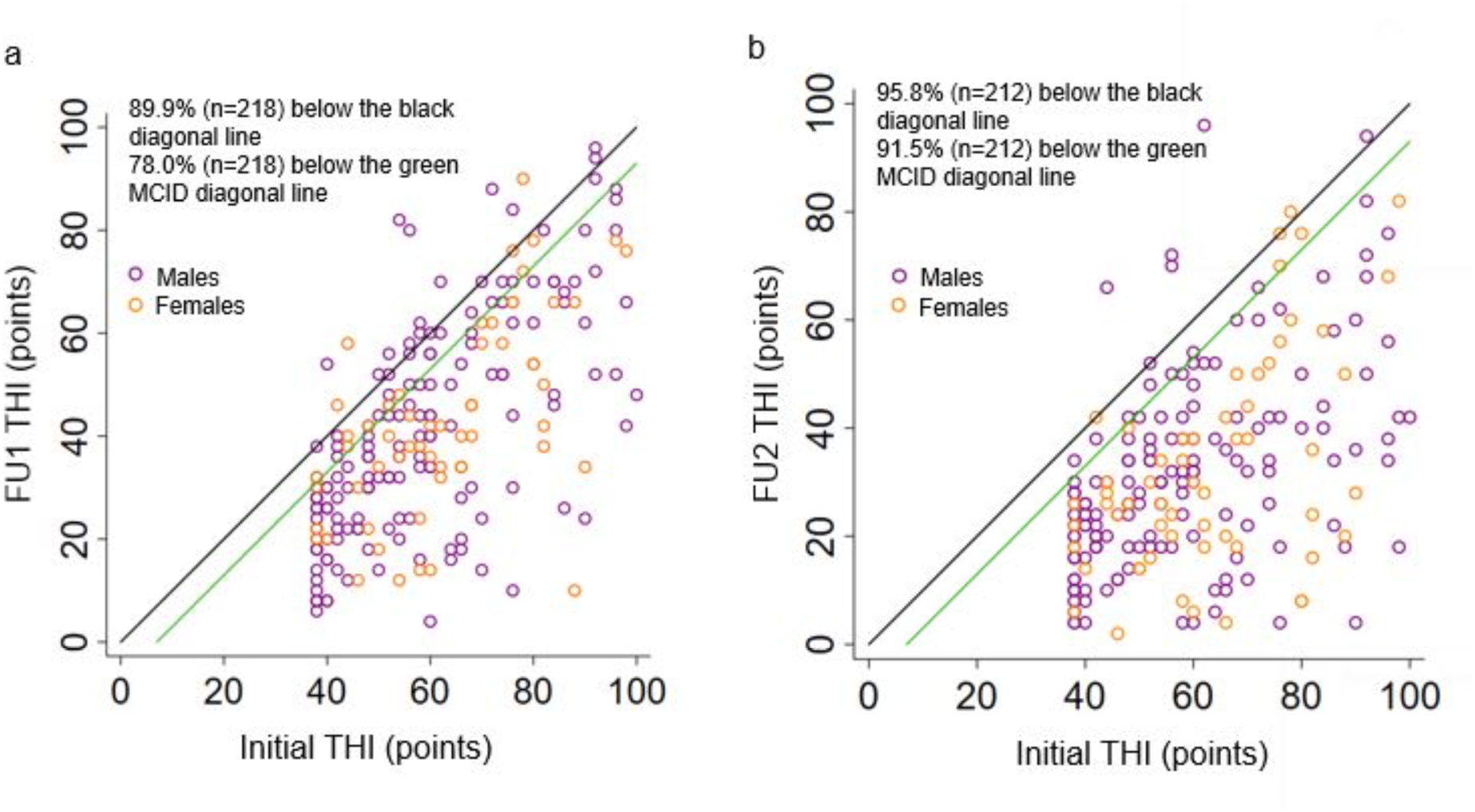
Scatter plots of THI scores of each patient at **(a)** initial assessment versus first follow-up (FU1), and **(b)** initial assessment versus second follow-up (FU2). Each circle representing each patient that is below the green line corresponds to an improvement by at least seven points on the THI scale based on the MCID, whereas each circle below the black line corresponds to any improvement in THI score.

**Supplementary Table 1.** Demographics and tinnitus characteristics for patients who did not attend their second follow-up (FU2) visit.

|  | Age (years) | Sex | THI at baseline (years) | Tinnitus duration at initial assessment (years) | Mean hearing loss at initial assessment (dB HL) |
| --- | --- | --- | --- | --- | --- |
| Patient 1 | 41-45 | Male | 76-80 | 21-25 | Right ear, 3.3<br>Left ear, 1.7 |
| Patient 2 | 51-55 | Male | 76-80 | 0-5 | Right ear, 5<br>Left ear, 1.7 |
| Patient 3 | 56-60 | Male | 81-85 | 0-5 | Right ear, 6.6<br>Left ear, 8.3 |
| Patient 4 | 51-55 | Female | 71-75 | 0-5 | Right ear, 10<br>Left ear, 13 |
| Patient 5 | 46-50 | Male | 51-55 | 6-10 | Right ear, 15<br>Left ear, 16.7 |
| Patient 6 | 66-70 | Male | 36-40 | 0-5 | Right ear, 20<br>Left ear, 20 |
| Patient 7 | 51-55 | Male | 76-80 | 16-20 | Right ear, 42<br>Left ear, 30 |
| Patient 8 | 61-65 | Male | 81-85 | 6-10 | Right ear, 15<br>Left ear, 62 |

Mean hearing loss at initial assessment was calculated using the average of 500 Hz, 1000 Hz and 2000 Hz for each ear. THI: Tinnitus Handicap Inventory; dB HL: decibel hearing level.

**Supplementary Table 2.** Tinnitus Handicap Inventory (THI) scores at initial assessment and first follow-up (FU1) visit for patients who did not attend their second follow-up (FU2) visit.

|  | **THI at initial assessment** | **THI at first follow-up (FU1)** | **Change in THI score from Initial assessment to first follow-up (FU1)** |
| --- | --- | --- | --- |
| Patient 1 | 76-80 | - | - |
| Patient 2 | 76-80 | 41-45 | -32 |
| Patient 3 | 81-85 | 66-70 | -14 |
| Patient 4 | 71-75 | - | - |
| Patient 5 | 51-55 | 41-45 | -10 |
| Patient 6 | 36-40 | 51-55 | 14 |
| Patient 7 | 76-80 | 66-70 | -6 |
| Patient 8 | 81-85 | 76-80 | -2 |

